# UTTARAKHAND COVID-19 CASES AND DEATHS: COMPARING THE DEMOGRAPHICS AND SUGGESTIVE STRATEGY TO PREVENT SIMILAR PANDEMICS IN FUTURE

**DOI:** 10.1101/2021.09.03.21263064

**Authors:** Gaurav Joshi, Akshara Pande, Omdeep Gupta, Anoop Nautiyal, Sanjay Jasola, Prashant Gahtori

**Affiliations:** School of Pharmacy, Graphic Era Hill University, Dehradun-248002, India; Department of Computer Science, Graphic Era Hill University, Dehradun-248002, India; Department of Management, Graphic Era Hill University, Dehradun-248002, India; SDC Foundation 69, Vasant Vihar Dehradun-248006, India; Vice Chancellor Office, Graphic Era Hill University, Dehradun-248002, India

**Keywords:** Covid-19, SARS-CoV-2, Uttarakhand, Infections, Healthcare, Mitigation policies

## Abstract

Coronavirus disease 19 (Covid-19) is causing a dramatic impact on human life worldwide. As of June 11 2021, later one has attributed more than 174 million confirmed cases and over 3.5 million deaths globally. Nonetheless, a World Bank Group flagship report features Covid-19 induced global crisis as the strongest post-recession since World WarII. Currently, all approved therapeutics or vaccines are strictly allowed for emergency use. Hence, in the absence of pharmaceutical interventions, it is vital to analyze data set covering the growth rates of positive human cases, number of recoveries, other factors, and future strategies to manage the growth of fatal Covid-19 effectively. The Uttarakhand state of India is snuggled in the lap of the Himalayas and occupies more people than Israel, Switzerland, Hong Kong, etc. This study analyzed state Covid-19 data, fetched from an authenticated government repository using Python 3.9 from April 1, 2020, to February 28, 2021. In the first wave, plain areas of Uttarakhand covering the districts Dehradun, Haridwar, Nainital, and U. S. Nagar were severely affected and reported peak positive cases during the 21st – 26th week. Other hands, the hilly terrains of Uttarakhand districts, including Chamoli, Pauri Garhwal, and Rudraprayag, reported a high number of positive cases between the 30th and 31st week, and other hilly districts reported an increase in Covid-19 cases during the 34th to 38th week. The highest recovery rate was attributed to the hilly district Rudraprayag. The analysis also revealed that a very high doubling rate was seen during the last week of May to the first week of Jun 2020. At last, based on this blueprint, we have suggested 6-points solutions for preventing the next pandemic.

## 1. Introduction

Severe Acute Respiratory Syndrome Coronavirus-2 (SARS-CoV-2) is an “evil genius” that invades the host cell like a lamb, tearing the organ system, resulting in death and stalling the world economy causing widespread unemployment, poverty, and hunger^1^. SARS-CoV-2 is responsible for Covid-19 diseases and the seventh coronavirus strain overall infecting *Homo sapiens*^2^. The situation is even critical due to non-availability of approved therapeutics against SARS-CoV-2^3^. Above all, the invasiveness of SARS-CoV-2 is further influenced by numerous intrinsic and extrinsic factors^4^. An analysis of Covid-19 impact on the country or multiple states may lead to a different picture considering the different timelines of the first case, infection rates, mortality, and many other parameters. Further measures took by different state governments might also vary from state to state and may differently affect the outcome of the disease. Thus, addressing each state will enable the Government to follow innovative measures to curb the pandemic growth with the limited number of resources they already possess. Although, all Indian states have foreseen the distress of Covid-19 in the year 2020 and followed by the second wave that just emerged during the writing of this manuscript. To foresee the actual effect of these underlying factors in Covid-19 cases and death, we proposed a statistical analysis in the ‘Uttarakhand’ state based on positivity, recovery, and death rate and comparing it with numerous intrinsic and extrinsic factors to overlay a clear impact of the current pandemic in the state.

In the northern states of India, Uttarakhand is nestled in the lap of the Himalayas and crossed by the sacred Ganges River^5^. The Indian Population Census 2011 proposed that current population of state is about 11.9 million and has become home to more people than countries like Israel, Switzerland, Hong Kong, etc. The rugged terrains of Uttarakhand, with crippled health care infrastructure, have further worsened the pandemic. The recent surge of 446 COVID-19 cases in a day and 23 deaths as of Sunday, 06-Jun-2021, according to Uttarakhand’s Government Covid19 Health Bulletin, even after imposing a state lockdown from 11-May-2021. Overall, Uttarakhand reports a total of 334,024 confirmed cases and 66,699 deaths during the writing of manuscript. Here, the state capital and district Dehradun all alone reported a total of 121 cases. Other 12 districts Haridwar, Pithoragarh, Tehri Garhwal, Udham Singh Nagar, Nainital, Uttarkashi, Chamoli, Pauri Garhwal, Rudraprayag, Almora, Bageshwar, and Champawat, reported 67, 61, 54, 26, 25, 23, 23, 20, 09, 07, 06, and 04 cases respectively. The driving motive behind current work is to correlate the underlying factors and improve preparedness if such a pandemic devastates the state in the future. Moreover, similar studies may assist other states and country(s) to re-evaluate not only the health care infrastructure but also ensure to relieve the economic burdens from its citizens to get away fromthe ongoing and similar pandemic in the future.

## 2. Material and Methods

Python 3.9 programming language was used to build an epidemic model, and all Covid-19 data was retrieved from authenticated and free to use resource data from Department of Medical Health and Family Welfare, Uttarakhand Government for about 12 months w.e.f. 15-Mar-2020 to 28-Feb-2021. The epidemic area in Uttarakhand was divided into thirteen parts covering all districts Almora, Bageshwar, Chamoli, Champawat, Dehradun, Haridwar, Nainital, Pauri Garhwal, Pithoragarh, Rudraprayag, Tehri Garhwal, U. S. Nagar, and Uttarkashi. The methodology consists of three steps, (i) Data collection, (ii) Data Conversion, and (iii) Information Extraction, as reported below.

### 2.1. Data Collection

All authenticated and free to use resource data is retrieved from Uttarakhand Government Covid-19 health bulletin for 12 months from 15-Mar-2020 to 28-Feb-2021 (https://health.uk.gov.in/pages/display/140-novel-corona-virus-guidelines-and-advisory-).

### 2.2. Data Conversion

We used intelligent text recognition and a logical structure engine to recognize words, lines, paragraphs, and the reading order in reported PDF documents. Python command-line utility pdf2txt.py was used to extract text contents and stored them in CSVfile format.

### 2.3. Information Extraction

All.csv files were converted to .xls format for analytical purposes. The final excel file consists of twelve fields for all 13 districts Almora, Bageshwar, Chamoli, Champawat, Dehradun, Haridwar, Nainital, Pauri Garhwal, Pithoragarh, Rudraprayag, Tehri Garhwal, U. S. Nagar, and Uttarkashi, for analytical purpose.

## 3. Results and Discussion

In this study, we analyzed first time the impact of Covid-19 on the Uttarakhand state. Here, the epidemic area is grouped in 13 different districts of Uttarakhand state. The first case was recorded on March 15, 2020 in this region. Hence, we collected the data from March 15, 2020, to February 28, 2021 from reliable data source Uttarakhand’s Government health portal. Firstly, the data was generated based on district demography including population, sex ratio, and density, and secondly, based on Covid-19 reported data. We further segregated data month-wise, considering the samples tested, positive cases, recovered cases, and mortality among 13 districts (**Table 1**).

**Table 1.** Compilation of essential parameters from the retrieved Covid-19 data and essential demographic features of 13 districts of Uttarakhand

| District | Population | Area<br>(per sq<br>km) | Population<br>Density | Month-Year | Sample<br>Tested | Positives | Recovered<br>cases | Deceased | %Positives/ %<br>Infection rate | % Recovered<br>cases | % Deceased |
| --- | --- | --- | --- | --- | --- | --- | --- | --- | --- | --- | --- |
| Almora | 622,506 | 3090 | 198 | March 2020 | 0 | 0 | 0 | 0 | 3.85 | 94.89 | 0.77 |
|  |  |  |  | April 2020 | 65 | 1 | 1 | 0 |  |  |  |
|  |  |  |  | May 2020 | 277 | 62 | 2 | 0 |  |  |  |
|  |  |  |  | June 2020 | 2780 | 113 | 150 | 2 |  |  |  |
|  |  |  |  | July 2020 | 7478 | 126 | 60 | 0 |  |  |  |
|  |  |  |  | August 2020 | 12424 | 262 | 172 | 0 |  |  |  |
|  |  |  |  | September 2020 | 21381 | 952 | 827 | 6 |  |  |  |
|  |  |  |  | October 2020 | 15492 | 335 | 491 | 1 |  |  |  |
|  |  |  |  | November 2020 | 8973 | 495 | 382 | 5 |  |  |  |
|  |  |  |  | December 2020 | 11401 | 716 | 670 | 10 |  |  |  |
|  |  |  |  | January 2021 | 4130 | 187 | 254 | 1 |  |  |  |
|  |  |  |  | February 2021 | 0 | 0 | 74 | 0 |  |  |  |
| Bageshwar | 259,898 | 2310 | 116 | March 2020 | 0 | 0 | 0 | 0 | 0.28 | 96.69 | 1.17 |
|  |  |  |  | April 2020 | 17 | 0 | 0 | 0 |  |  |  |
|  |  |  |  | May 2020 | 328 | 16 | 0 | 0 |  |  |  |
|  |  |  |  | June 2020 | 1126 | 67 | 66 | 1 |  |  |  |
|  |  |  |  | July 2020 | 2579 | 19 | 28 | 0 |  |  |  |
|  |  |  |  | August 2020 | 6461 | 172 | 93 | 0 |  |  |  |
|  |  |  |  | September 2020 | 9161 | 358 | 333 | 3 |  |  |  |
|  |  |  |  | October 2020 | 13162 | 301 | 277 | 4 |  |  |  |
|  |  |  |  | November 2020 | 8308 | 234 | 230 | 1 |  |  |  |
|  |  |  |  | December 2020 | 8033 | 333 | 300 | 9 |  |  |  |
|  |  |  |  | January 2021 | 6451 | 39 | 70 | 0 |  |  |  |
|  |  |  |  | February 2021 | 0 | 0 | 91 | 0 |  |  |  |
| Chamoli | 391,605 | 7692 | 49 | March 2020 | 0 | 0 | 3 | 0 | 3.86 | 96.87 | 0.43 |
|  |  |  |  | April 2020 | 10 | 0 | 0 | 0 |  |  |  |
|  |  |  |  | May 2020 | 617 | 13 | 0 | 0 |  |  |  |
|  |  |  |  | June 2020 | 2015 | 61 | 62 | 0 |  |  |  |
|  |  |  |  | July 2020 | 3428 | 18 | 20 | 0 |  |  |  |
|  |  |  |  | August 2020 | 10122 | 262 | 154 | 0 |  |  |  |
|  |  |  |  | September 2020 | 15832 | 688 | 508 | 0 |  |  |  |
|  |  |  |  | October 2020 | 15129 | 758 | 832 | 1 |  |  |  |
|  |  |  |  | November 2020 | 13727 | 874 | 770 | 11 |  |  |  |
|  |  |  |  | December 2020 | 20404 | 703 | 831 | 3 |  |  |  |
|  |  |  |  | January 2021 | 9093 | 82 | 170 | 0 |  |  |  |
|  |  |  |  | February 2021 | 0 | 28 | 28 | 0 |  |  |  |
| Champawat | 259,648 | 1781 | 147 | March 2020 | 0 | 0 | 0 | 0 | 1.92 | 97.86 | 0.49 |
|  |  |  |  | April 2020 | 26 | 0 | 0 | 0 |  |  |  |
|  |  |  |  | May 2020 | 437 | 12 | 0 | 0 |  |  |  |
|  |  |  |  | June 2020 | 850 | 47 | 43 | 1 |  |  |  |
|  |  |  |  | July 2020 | 5281 | 64 | 22 | 0 |  |  |  |
|  |  |  |  | August 2020 | 9735 | 184 | 145 | 1 |  |  |  |
|  |  |  |  | September 2020 | 18196 | 537 | 357 | 2 |  |  |  |
|  |  |  |  | October 2020 | 13430 | 301 | 460 | 2 |  |  |  |
|  |  |  |  | November 2020 | 15175 | 244 | 141 | 1 |  |  |  |
|  |  |  |  | December 2020 | 20282 | 333 | 349 | 1 |  |  |  |
|  |  |  |  | January 2021 | 11633 | 96 | 200 | 1 |  |  |  |
|  |  |  |  | February 2021 | 0 | 7 | 69 | 0 |  |  |  |
| Dehradun | 1,696,694 | 3088 | 550 | March 2020 | 60 | 6 | 10 | 0 | 7.10 | 94.93 | 3.24 |
|  |  |  |  | April 2020 | 1810 | 31 | 17 | 0 |  |  |  |
|  |  |  |  | May 2020 | 6401 | 203 | 21 | 3 |  |  |  |
|  |  |  |  | June 2020 | 11190 | 458 | 513 | 21 |  |  |  |
|  |  |  |  | July 2020 | 16018 | 967 | 616 | 20 |  |  |  |
|  |  |  |  | August 2020 | 28950 | 2426 | 1556 | 87 |  |  |  |
|  |  |  |  | September 2020 | 41284 | 9118 | 7259 | 163 |  |  |  |
|  |  |  |  | October 2020 | 51240 | 4047 | 5744 | 295 |  |  |  |
|  |  |  |  | November 2020 | 63741 | 4123 | 3328 | 94 |  |  |  |
|  |  |  |  | December 2020 | 105923 | 5900 | 5505 | 162 |  |  |  |
|  |  |  |  | January 2021 | 91786 | 2020 | 3166 | 88 |  |  |  |
|  |  |  |  | February 2021 | 0 | 425 | 483 | 30 |  |  |  |
| Haridwar | 1,890,422 | 2360 | 801 | March 2020 | 16 | 0 | 0 | 0 | 3.84 | 96.67 | 1.11 |
|  |  |  |  | April 2020 | 997 | 7 | 5 | 0 |  |  |  |
|  |  |  |  | May 2020 | 4307 | 61 | 2 | 0 |  |  |  |
|  |  |  |  | June 2020 | 4803 | 245 | 229 | 0 |  |  |  |
|  |  |  |  | July 2020 | 14420 | 1103 | 279 | 2 |  |  |  |
|  |  |  |  | August 2020 | 39740 | 3302 | 2988 | 43 |  |  |  |
|  |  |  |  | September 2020 | 53107 | 4643 | 4241 | 47 |  |  |  |
|  |  |  |  | October 2020 | 54762 | 1637 | 2637 | 29 |  |  |  |
|  |  |  |  | November 2020 | 63452 | 1089 | 881 | 14 |  |  |  |
|  |  |  |  | December 2020 | 67270 | 1294 | 1467 | 12 |  |  |  |
|  |  |  |  | January 2021 | 66384 | 606 | 765 | 5 |  |  |  |
|  |  |  |  | February 2021 | 0 | 201 | 221 | 6 |  |  |  |
| Nainital | 954,605 | 3853 | 225 | March 2020 | 11 | 1 | 7 | 0 | 6.20 | 97.08 | 1.87 |
|  |  |  |  | April 2020 | 593 | 2 | 8 | 0 |  |  |  |
|  |  |  |  | May 2020 | 3322 | 250 | 15 | 1 |  |  |  |
|  |  |  |  | June 2020 | 3598 | 214 | 317 | 2 |  |  |  |
|  |  |  |  | July 2020 | 8544 | 630 | 329 | 12 |  |  |  |
|  |  |  |  | August 2020 | 15479 | 1592 | 1242 | 37 |  |  |  |
|  |  |  |  | September 2020 | 25422 | 3212 | 2943 | 60 |  |  |  |
|  |  |  |  | October 2020 | 33560 | 1421 | 1899 | 33 |  |  |  |
|  |  |  |  | November 2020 | 35611 | 1210 | 1133 | 24 |  |  |  |
|  |  |  |  | December 2020 | 41472 | 2785 | 2216 | 37 |  |  |  |
|  |  |  |  | January 2021 | 36536 | 1221 | 1930 | 24 |  |  |  |
|  |  |  |  | February 2021 | 0 | 112 | 242 | 7 |  |  |  |
| Pauri Garhwal | 687,271 | 5438 | 129 | March 2020 | 2 | 0 | 0 | 0 | 3.71 | 98.49 | 1.16 |
|  |  |  |  | April 2020 | 21 | 1 | 1 | 0 |  |  |  |
|  |  |  |  | May 2020 | 475 | 33 | 2 | 1 |  |  |  |
|  |  |  |  | June 2020 | 3696 | 108 | 88 | 2 |  |  |  |
|  |  |  |  | July 2020 | 5888 | 68 | 88 | 1 |  |  |  |
|  |  |  |  | August 2020 | 13388 | 287 | 172 | 1 |  |  |  |
|  |  |  |  | September 2020 | 14056 | 1482 | 940 | 15 |  |  |  |
|  |  |  |  | October 2020 | 35766 | 991 | 1139 | 9 |  |  |  |
|  |  |  |  | November 2020 | 25218 | 1260 | 1182 | 15 |  |  |  |
|  |  |  |  | December 2020 | 23276 | 759 | 1140 | 13 |  |  |  |
|  |  |  |  | January 2021 | 17019 | 166 | 291 | 1 |  |  |  |
|  |  |  |  | February 2021 | 0 | 0 | 34 | 2 |  |  |  |
| **Pithoragarh** | 483,439 | 7110 | 68 | March 2020 | 0 | 0 | 0 | 0 | 4.63 | 98.25 | 1.43 |
|  |  |  |  | April 2020 | 1 | 0 | 0 | 0 |  |  |  |
|  |  |  |  | May 2020 | 291 | 21 | 0 | 0 |  |  |  |
|  |  |  |  | June 2020 | 1452 | 44 | 52 | 0 |  |  |  |
|  |  |  |  | July 2020 | 5478 | 78 | 18 | 0 |  |  |  |
|  |  |  |  | August 2020 | 10535 | 196 | 143 | 2 |  |  |  |
|  |  |  |  | September 2020 | 14643 | 735 | 605 | 3 |  |  |  |
|  |  |  |  | October 2020 | 8041 | 396 | 443 | 1 |  |  |  |
|  |  |  |  | November 2020 | 9919 | 775 | 553 | 16 |  |  |  |
|  |  |  |  | December 2020 | 15641 | 968 | 922 | 16 |  |  |  |
|  |  |  |  | January 2021 | 6668 | 132 | 495 | 7 |  |  |  |
|  |  |  |  | February 2021 | 0 | 17 | 72 | 3 |  |  |  |
| **Rudraprayag** | 242,285 | 1896 | 127 | March 2020 | 0 | 0 | 0 | 0 | 4.33 | 99.21 | 0.44 |
|  |  |  |  | April 2020 | 12 | 0 | 0 | 0 |  |  |  |
|  |  |  |  | May 2020 | 424 | 6 | 0 | 0 |  |  |  |
|  |  |  |  | June 2020 | 1221 | 60 | 59 | 1 |  |  |  |
|  |  |  |  | July 2020 | 3501 | 10 | 7 | 0 |  |  |  |
|  |  |  |  | August 2020 | 7696 | 142 | 96 | 0 |  |  |  |
|  |  |  |  | September 2020 | 9141 | 474 | 366 | 0 |  |  |  |
|  |  |  |  | October 2020 | 12025 | 587 | 450 | 1 |  |  |  |
|  |  |  |  | November 2020 | 5393 | 574 | 768 | 6 |  |  |  |
|  |  |  |  | December 2020 | 9188 | 347 | 365 | 2 |  |  |  |
|  |  |  |  | January 2021 | 3768 | 67 | 116 | 0 |  |  |  |
|  |  |  |  | February 2021 | 0 | 3 | 25 | 0 |  |  |  |
| **Tehri Garhwal** | 618,931 | 4085 | 170 | March 2020 | 0 | 0 | 0 | 0 | 4.03 | 95.16 | 0.38 |
|  |  |  |  | April 2020 | 31 | 0 | 0 | 0 |  |  |  |
|  |  |  |  | May 2020 | 1537 | 77 | 0 | 0 |  |  |  |
|  |  |  |  | June 2020 | 2452 | 339 | 383 | 2 |  |  |  |
|  |  |  |  | July 2020 | 5606 | 101 | 75 | 0 |  |  |  |
|  |  |  |  | August 2020 | 18801 | 632 | 369 | 0 |  |  |  |
|  |  |  |  | September 2020 | 21183 | 1178 | 1124 | 1 |  |  |  |
|  |  |  |  | October 2020 | 15165 | 780 | 873 | 2 |  |  |  |
|  |  |  |  | November 2020 | 9866 | 464 | 415 | 8 |  |  |  |
|  |  |  |  | December 2020 | 19401 | 553 | 493 | 3 |  |  |  |
|  |  |  |  | January 2021 | 11060 | 103 | 265 | 0 |  |  |  |
|  |  |  |  | February 2021 | 0 | 6 | 31 | 0 |  |  |  |
| US Nagar | 1,648,902 | 2912 | 648 | March 2020 | 0 | 0 | 0 | 0 | 3.43 | 97.69 | 1.01 |
|  |  |  |  | April 2020 | 977 | 7 | 4 | 0 |  |  |  |
|  |  |  |  | May 2020 | 2668 | 75 | 23 | 0 |  |  |  |
|  |  |  |  | June 2020 | 4612 | 177 | 120 | 3 |  |  |  |
|  |  |  |  | July 2020 | 15500 | 993 | 318 | 4 |  |  |  |
|  |  |  |  | August 2020 | 36334 | 2509 | 1788 | 17 |  |  |  |
|  |  |  |  | September 2020 | 51219 | 4674 | 5046 | 38 |  |  |  |
|  |  |  |  | October 2020 | 52205 | 1045 | 1713 | 31 |  |  |  |
|  |  |  |  | November 2020 | 49451 | 822 | 771 | 10 |  |  |  |
|  |  |  |  | December 2020 | 59285 | 799 | 930 | 6 |  |  |  |
|  |  |  |  | January 2021 | 64891 | 358 | 396 | 8 |  |  |  |
|  |  |  |  | February 2021 | 0 | 90 | 173 | 0 |  |  |  |
| Uttarkashi | 330,086 | 7951 | 41 | March 2020 | 0 | 0 | 0 | 0 | 3.16 | 96.20 | 0.45 |
|  |  |  |  | April 2020 | 85 | 0 | 0 | 0 |  |  |  |
|  |  |  |  | May 2020 | 489 | 21 | 1 | 0 |  |  |  |
|  |  |  |  | June 2020 | 1418 | 45 | 43 | 1 |  |  |  |
|  |  |  |  | July 2020 | 5368 | 125 | 77 | 0 |  |  |  |
|  |  |  |  | August 2020 | 16237 | 678 | 522 | 1 |  |  |  |
|  |  |  |  | September 2020 | 23484 | 1122 | 878 | 4 |  |  |  |
|  |  |  |  | October 2020 | 18108 | 729 | 930 | 3 |  |  |  |
|  |  |  |  | November 2020 | 15945 | 303 | 350 | 3 |  |  |  |
|  |  |  |  | December 2020 | 21582 | 635 | 491 | 4 |  |  |  |
|  |  |  |  | January 2021 | 17240 | 132 | 342 | 1 |  |  |  |
|  |  |  |  | February 2021 | 0 | 0 | 12 | 0 |  |  |  |

The cumulative analysis of data revealed (**Figure 1**) that during the course of analysis (April 11, 2020, to February 28, 2021), the State of Uttarakhand tested a total of 2.143295 million samples (about 34,224 tests per million populations). This figure is below the national average of 81,852 tests per million populations during the period of analysis. All tests were facilitated by 54 Government and 09 private laboratories, all approved by the central apex body, Indian Council of Medical Research (ICMR), New Delhi. Among the tested population, the region attained 97,021 positive cases, which are approximately 4.52 positives per 100 people tested. Further analysis suggested that different districts of the region faced Covid-19 waves unequivocally (**Figure 2A**). For instance, among 13 districts, Dehradun, with a total of 29,724 cases, was worst affected during the first COVID-19 wave. This was followed by Haridwar (14,188), Nainital (12,650), and U. S. Nagar (11,549). All these four districts have large areas in the plains and possess larger populations. In terms of percentage infection rate of samples tested district-wise, the highest positive cases was appeared in Dehradun (7.10%), followed by Nainital (6.20%) and Pithoragarh (4.63%). The lowest infection rates were displayed by district Champawat (1.92%), followed by Bageshwar (2.77%) and Uttarkashi (3.16%).

**Figure 1.**
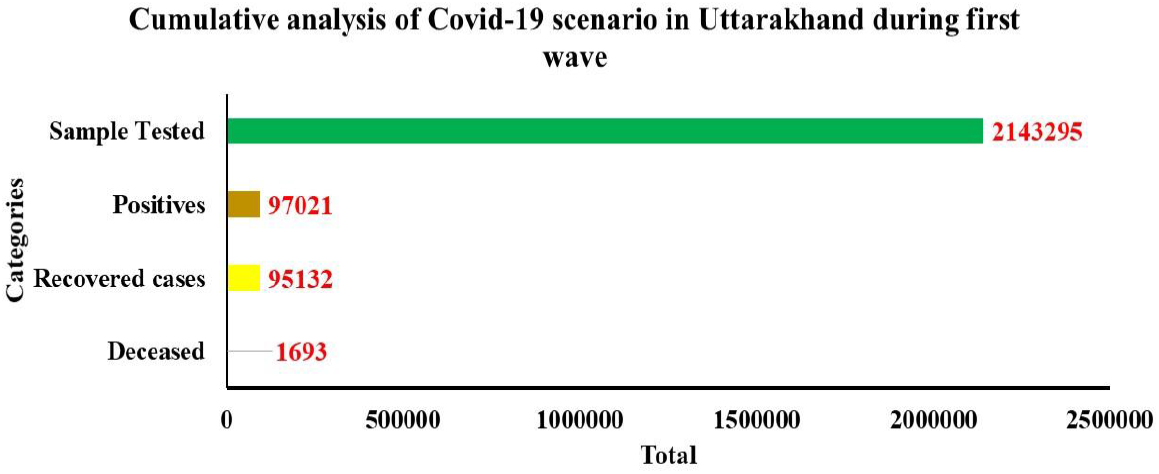
The bar graphs illustrate total testing done, recovered cases, and cumulative death in the state of Uttarakhandcompiled from April 2020 to February 2021

**Figure 2.**
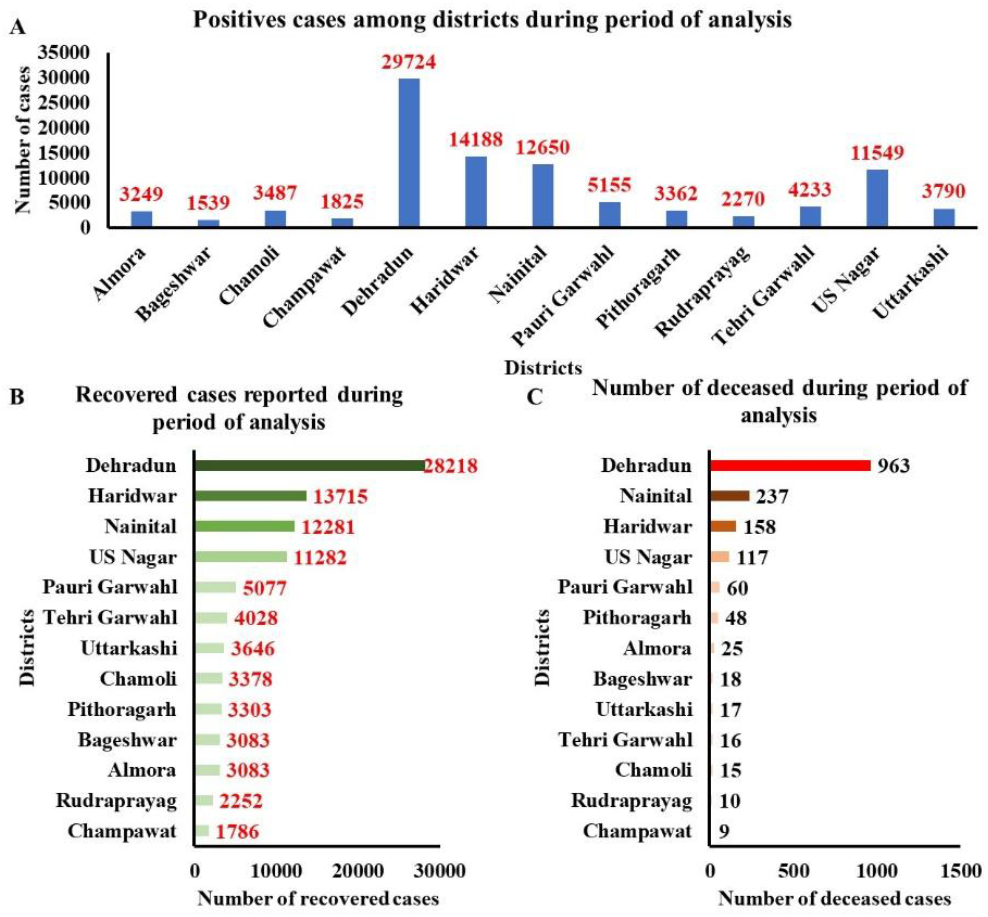
The bar graphs were compiled based on Covid-19 severity amongst 13 districts of Uttarakhand from April 2020 to February 2021. **A**.Covid-19 positives identified; **B**. Recovered cases and **C**. mortality among 13 districts of Uttarakhand.

In contrast, the major sign of relief was the fast recovery rate at par with the positivity cases. The data analyzed revealed (**Figure 2B**) that among 13 districts, Dehradun, with the highest number of positive cases, revealed the highest recovery (28,218) that accounted for a total 30.72% share of all other districts. This trend was followed by Haridwar (11,282), Nainital (12,281), and U.S. Nagar (13,715).A total of 1,645 people died in the state during the period of analysis. Here, the optimal number of deaths occurred in Dehradun (3.24 %) of tested positives. District Nainital and Pithoragarh were placed in second and third-position with percentage deaths 1.87% and 1.43%, respectively. The lowest deaths were displayed by district Tehri Garhwal (0.38%). The significant deaths were reported (**Figure 2C**) from districts of Dehradun (963 deaths), which was approximately accounted for 58.54% of total deaths that cumulatively occurred in the state. Nainital district recorded 237 (14.40%) deaths, followed by Haridwar and U. S. Nagar with 158 (9.60%) and 117 (7.11%) of total death, respectively.

The Government of Uttarakhand acted quickly by imposing restrictions and partial lockdown from last year March 18, 2020, till March 31, 2020. This included banning tourism entries, closing schools, cinema halls, places of mass gathering, Janta curfew, the National bio reserves, and thedevelopment of containment zones in different districts and high-risk areas. The lockdown was further extended to May 3, 2020, by the Uttarakhand state government, considering the surge in cases. The lockdown was slowly uplifted from May 4, 2020, under various phases considering the safety protocols. We analyzed the effect of Covid-19 restrictions that includes imposing lockdown, upliftment of lockdown in various phases on Covid-19 cases, recovered cases, and existing cases per week among 13 districts. The analysis revealed (**Figure 3**) that during the lockdown (1-4 weeks; April 11 to May 3, 2020), Covid-19 cases were strictly under control. However, the steep increase in Covid-19 cases was observed in 21^st^ – 26^th^ and 34^th^ to 38^th^ week. The Dehradun was worst affected (**Figure 3E**) during the 21^st^ – 26^th^ week period and reported an average of approximately 4316 cases. This was followed by Haridwar (**Figure 3F**) (2482), Nainital (1490) (**Figure 3G**),and US Nagar (1453) (**Figure 3M**) during the 21^st^ – 26^th^week. However, there was a steep decline in cases in-between 34^th^ to 38^th^ week in districts like Dehradun (2932), U. S. Nagar (523), and Haridwar (904). Moreover, the hilly terrains of Uttarakhand, including the district of Chamoli (**Figure 3C**), Pauri Garhwal (**Figure 3H**), and Rudraprayag (**Figure 3J**), reported a high number of positive cases in the 30^th^ and 31^st^ week.

**Figure 3(A-M).**
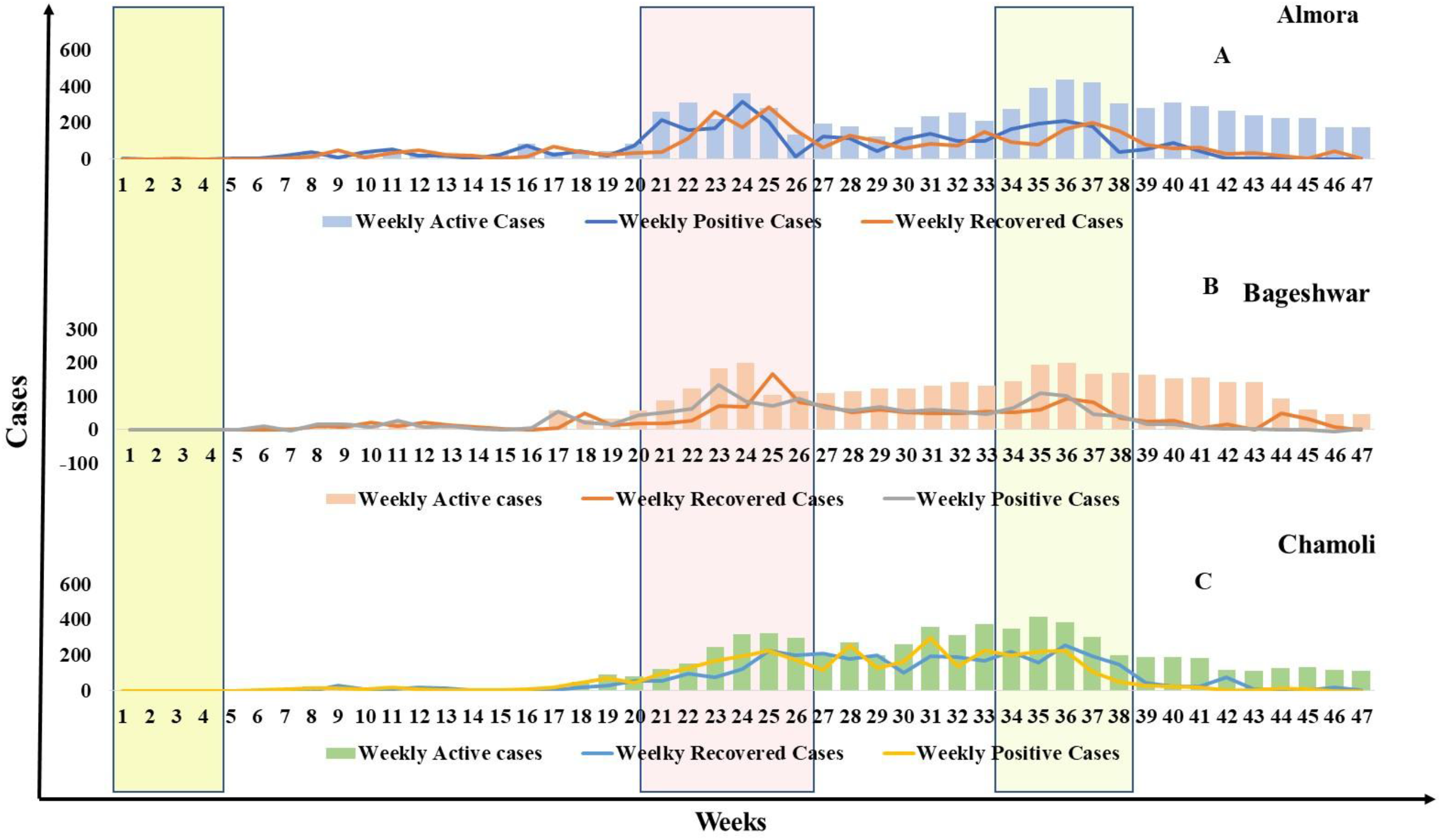
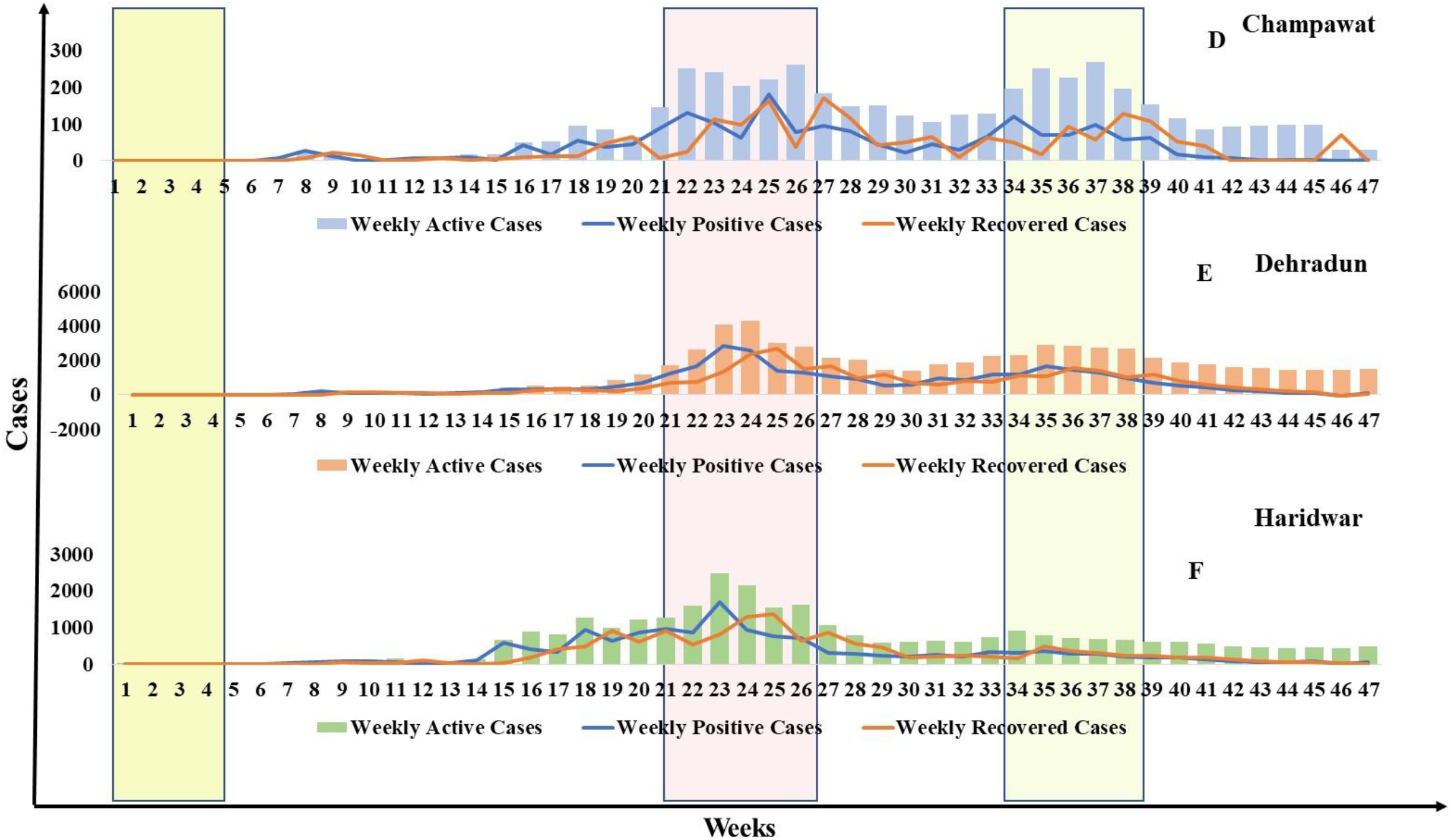
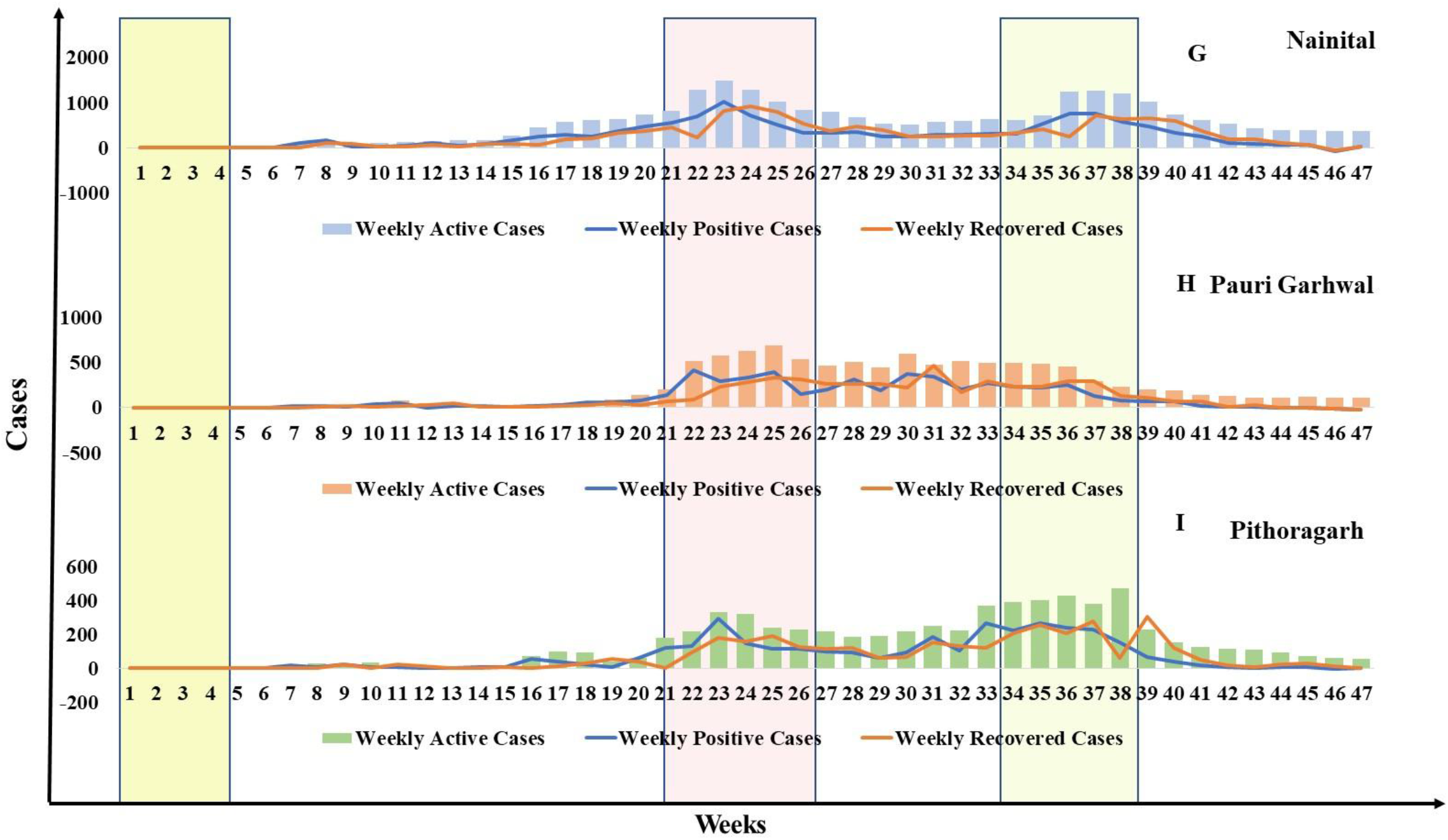
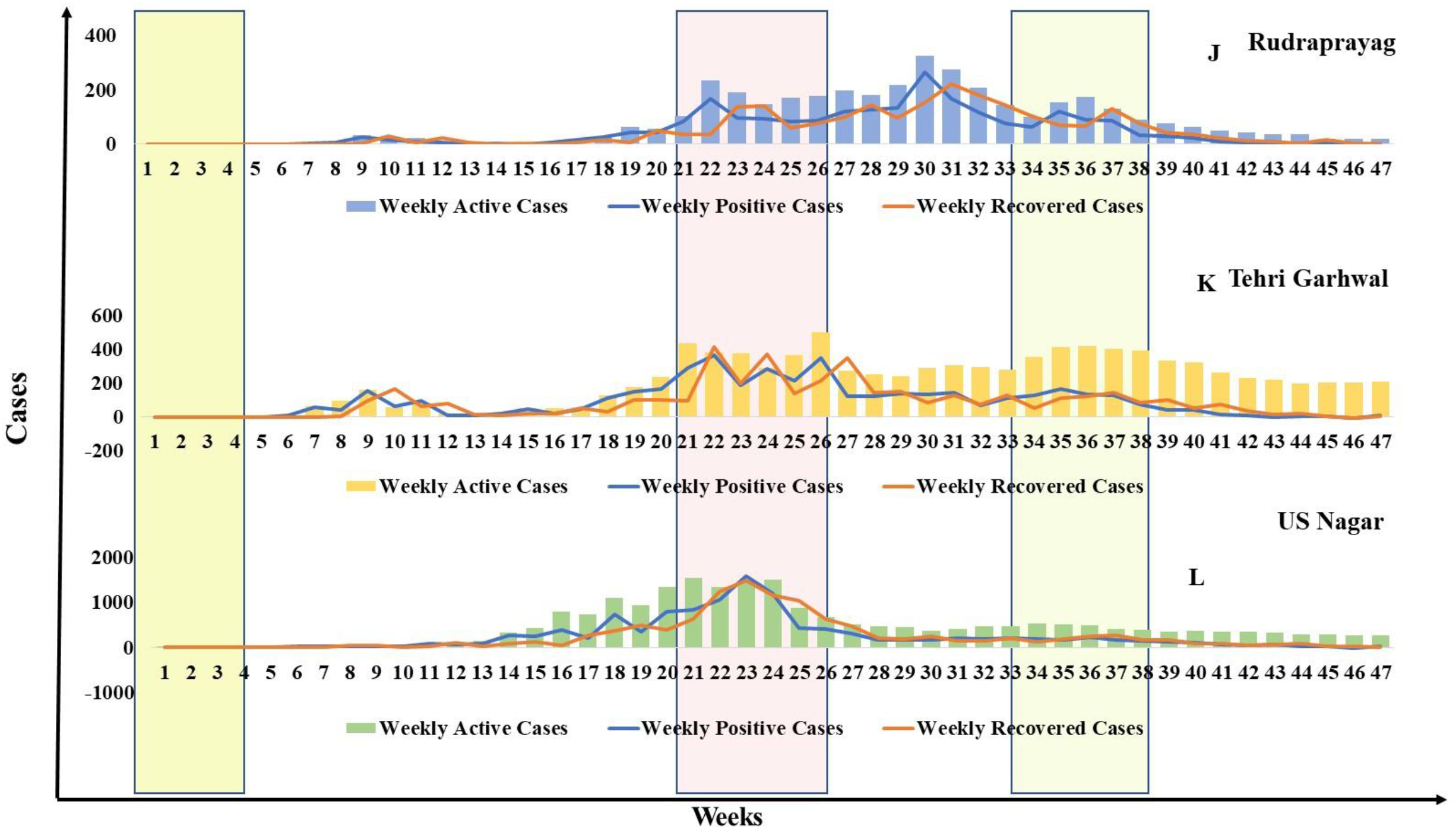
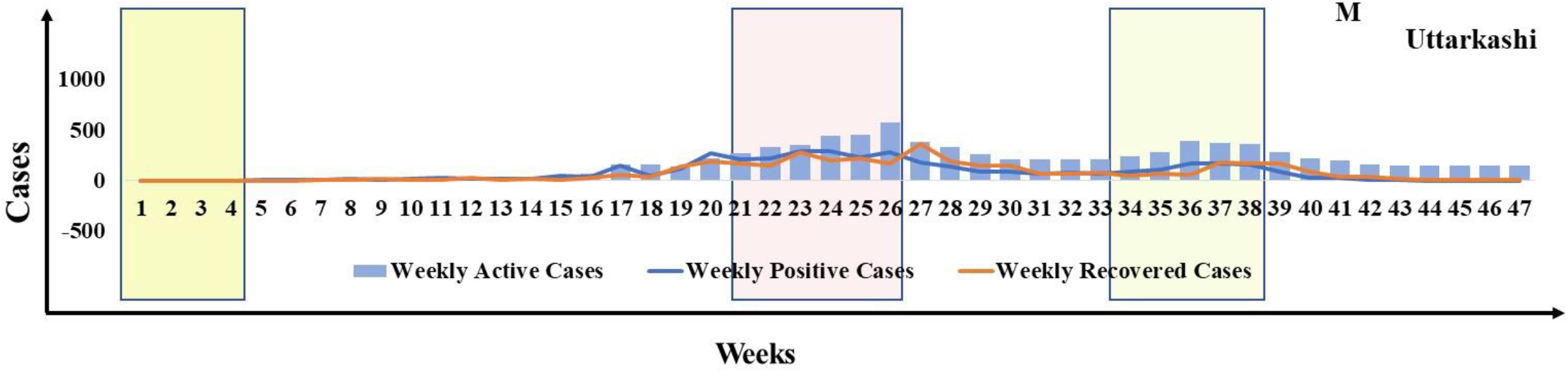
Bar graphs reveal the effect of Covid-19 restrictions on the outcome of Covid-19 cases, recovered cases, and existing cases per week among 13 districts.The graph revealed that the positive cases were directly correlated with recovered cases.

In addition, almost every hilly district of Uttarakhand reported an increase in Covid-19 cases during the 34^th^ to 38^th^ week. The case reported during this span was maximum compared to their average cases from 1^st^ to 47^th^ week (**Figure 4**).

**Figure 4.**
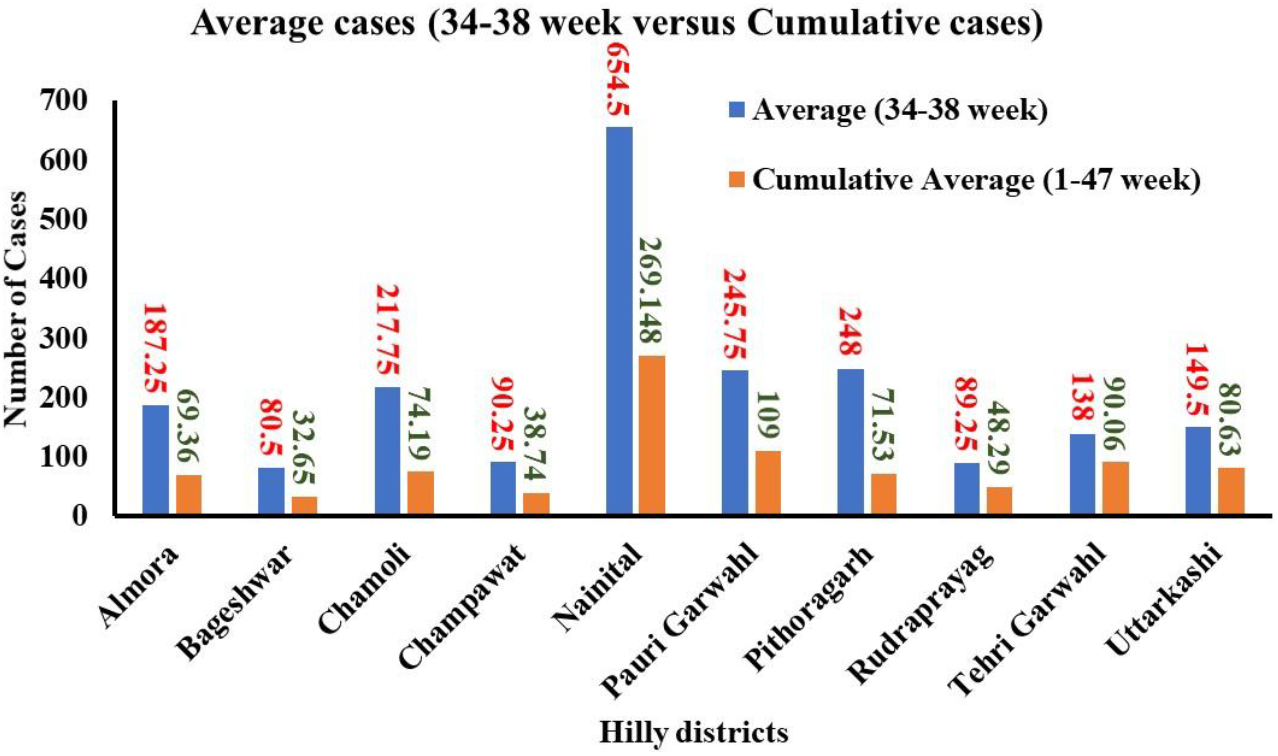
Bar graphs suggesting a high impact of Covid-19 in infecting the population of 13 districts of Uttarakhand during the 34^th^ to 38^th^ week concerning total weeks of analysis, i.e., 1^st^ to 47^th^ week

Furthermore, the analysis revealed that Uttarakhandhad attaineda high recovery of 96.41% of total positive COVID-19 cases. Here, the district Rudraprayag conferred with the highest recovery (99.21%), followed by Pauri Garhwal (98.49%) and Pithoragarh (98.25%) (**Figure 5**). Despite the high recovery shown by district Almora (94.89%) during the pandemic time, it appears the lowest than other districts.

**Figure 5.**
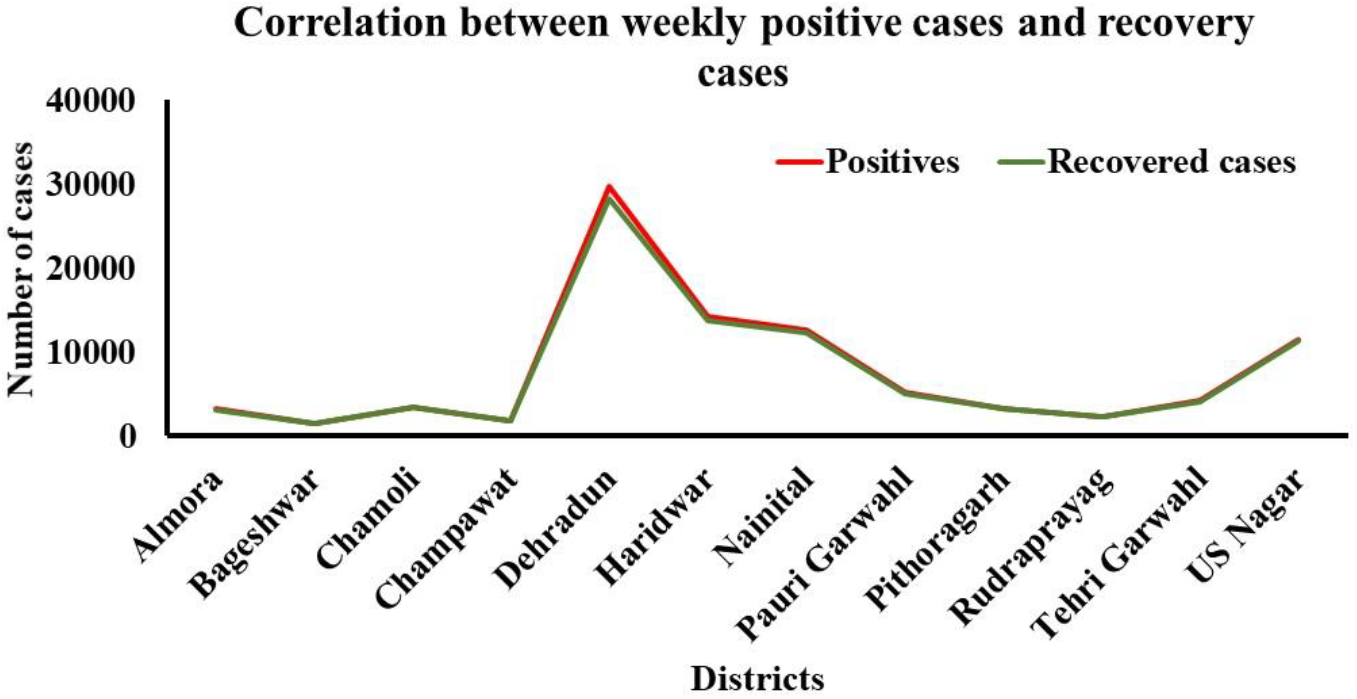
Line diagram reveals a high correlation between Covid-19 positive cases with their recovery rates among 13 districts of Uttarakhand

This is further marked with a doubling interval of Covid-19 cases in different districts of Uttarakhand. A doubling rate portrays the number of days in which virus growth turns 2-folds. Here, the outcomes are depicted in a table wherethe intensity of the doubling rate (along with date) is graded by color (**See Tables 2A and 2B**). The doubling rate of 2 - 2.9 is graded as ‘moderate,’ 3 - 4.9 is graded as ‘high,’ and 5 - 7 is graded as ‘very high.’ The analysis revealed that Uttarakhand had attained maximum growth of the virus from 23-May-2020 to 06-Jun-2020 with the lowest doubling rate at district Tehri Garhwal (1.93) last week of May 2020. The U. S. Nagar has seen the lowest virus growth rate with a very high doubling rate of 5.54 as of 25-May-2020, followed by a high doubling rate at Dehradun (3.84) as on 06-Jun-2020,Bageshwar (3.50) as on 26-May-2020, Rudraprayag (3.27) as on 06-Jun-2020 and Haridwar (3.07)) as on 31-May-2020. Overall, Tehri Garhwal was found to worst affected with the highest virus replication as deduced by the analysis as of May 30, 2020.

**Table 2A-B.**
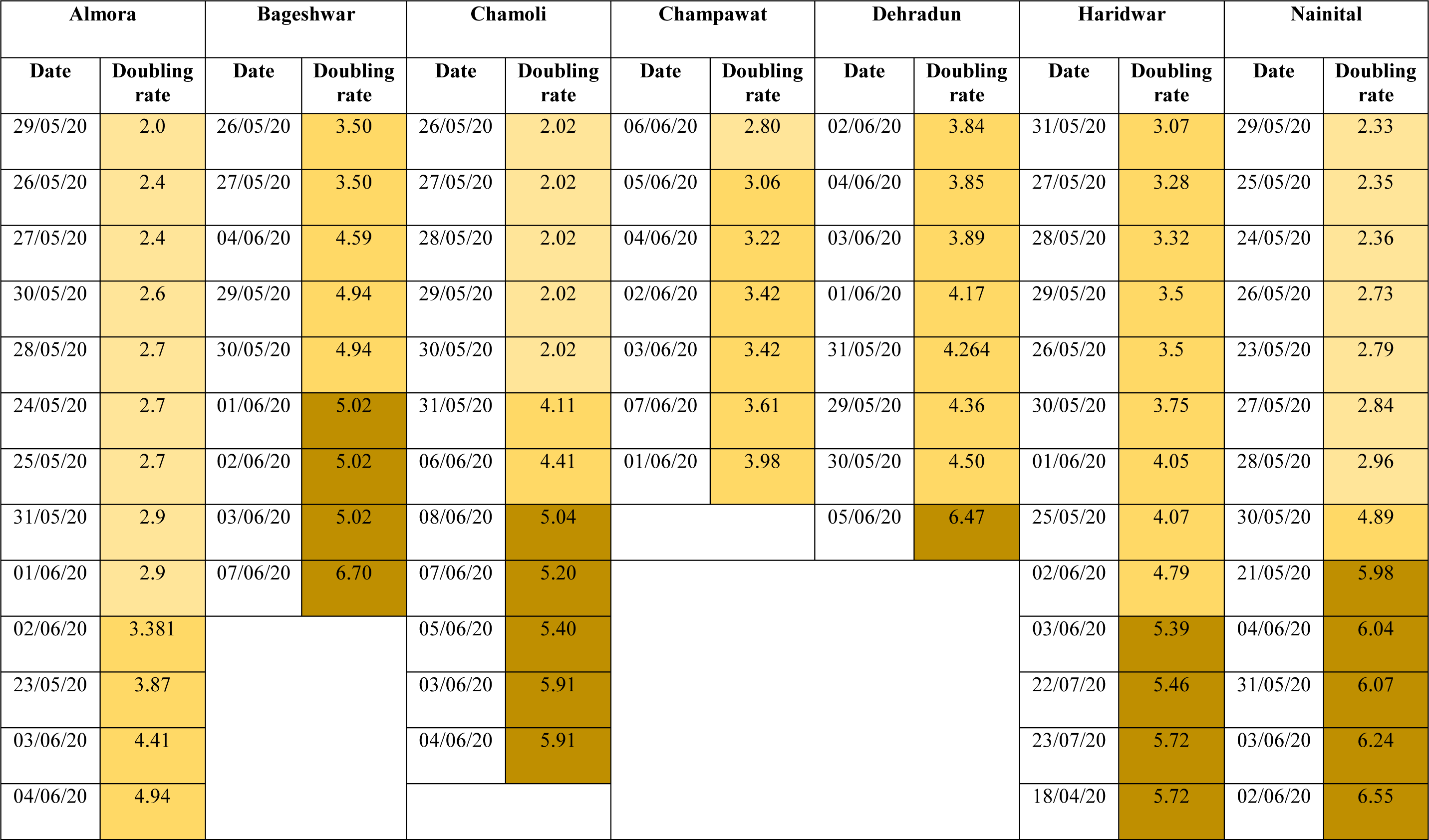
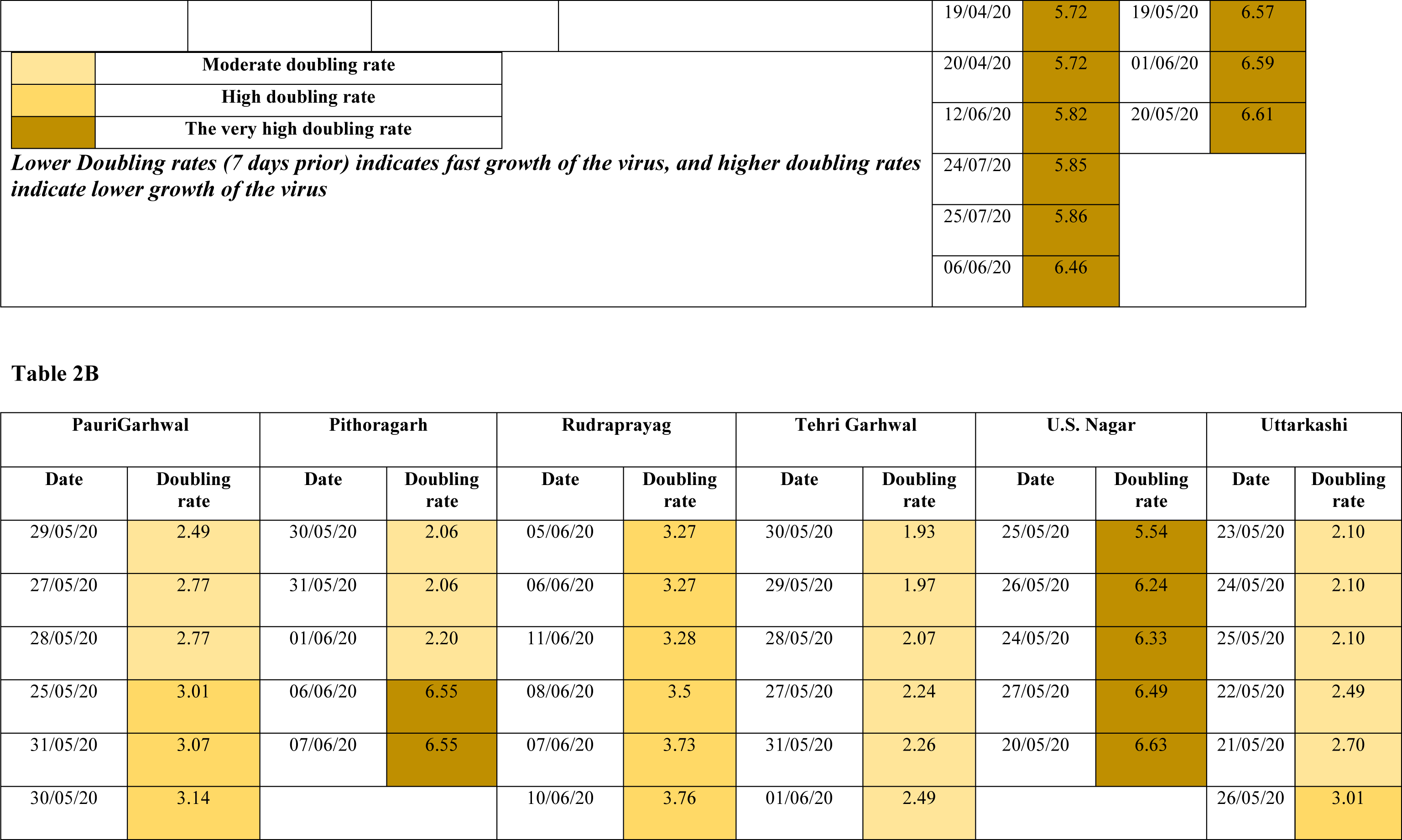
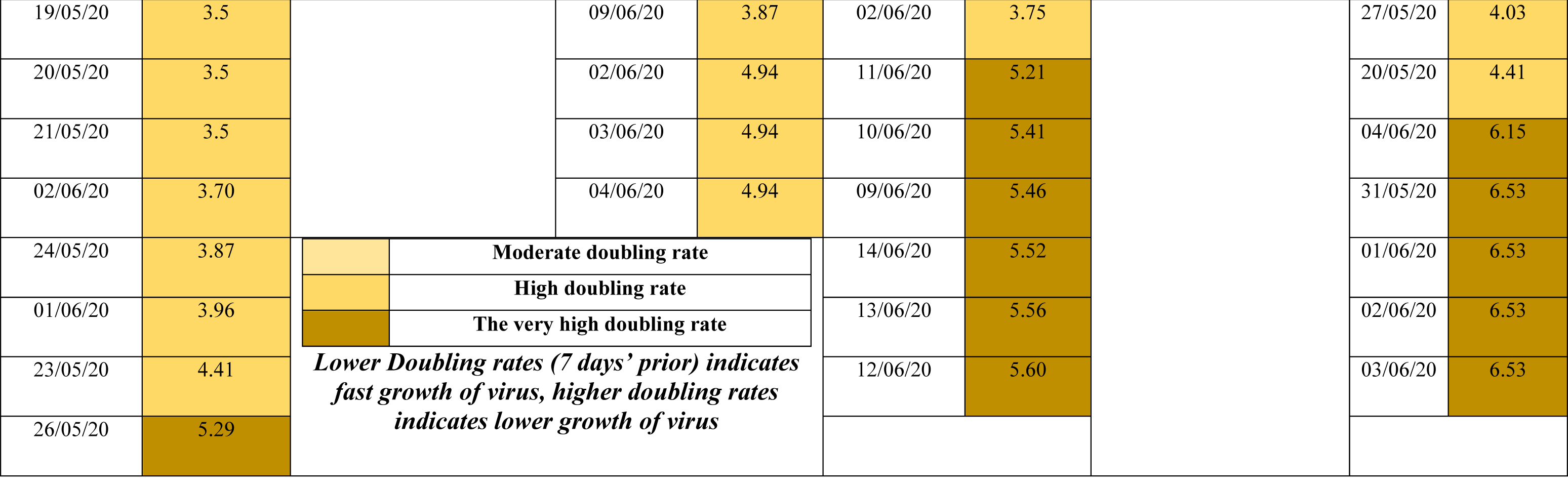
The table accounts for the doubling rates of Covid-19 in 13 districts of Uttarakhand during the period of analysis. The intensity of the doubling rate (along with date) is graded by color. The doubling rate of 2 - 2.9 is graded as ‘moderate,’ 3 - 4.9 is graded as ‘high,’ and 5 - 7 is graded as ‘very high.’

Next, we thought to compare the positivity rate of different districts of Uttarakhand based on their population and density. As the Census is due for Uttarakhand in 2021, we took the recent original data of 2011 when the last census study was last made^6^. However, for ease of correctness and to improve data significance in the gap of 10 years from the last Census, we also took the estimated population of Uttarakhand till the year 2021^7^. The analysis revealed (**Figure 6**) that population was one factor that portrayed a significantly high correlation (avg. r^2^: 0.83).

**Figure 6.**
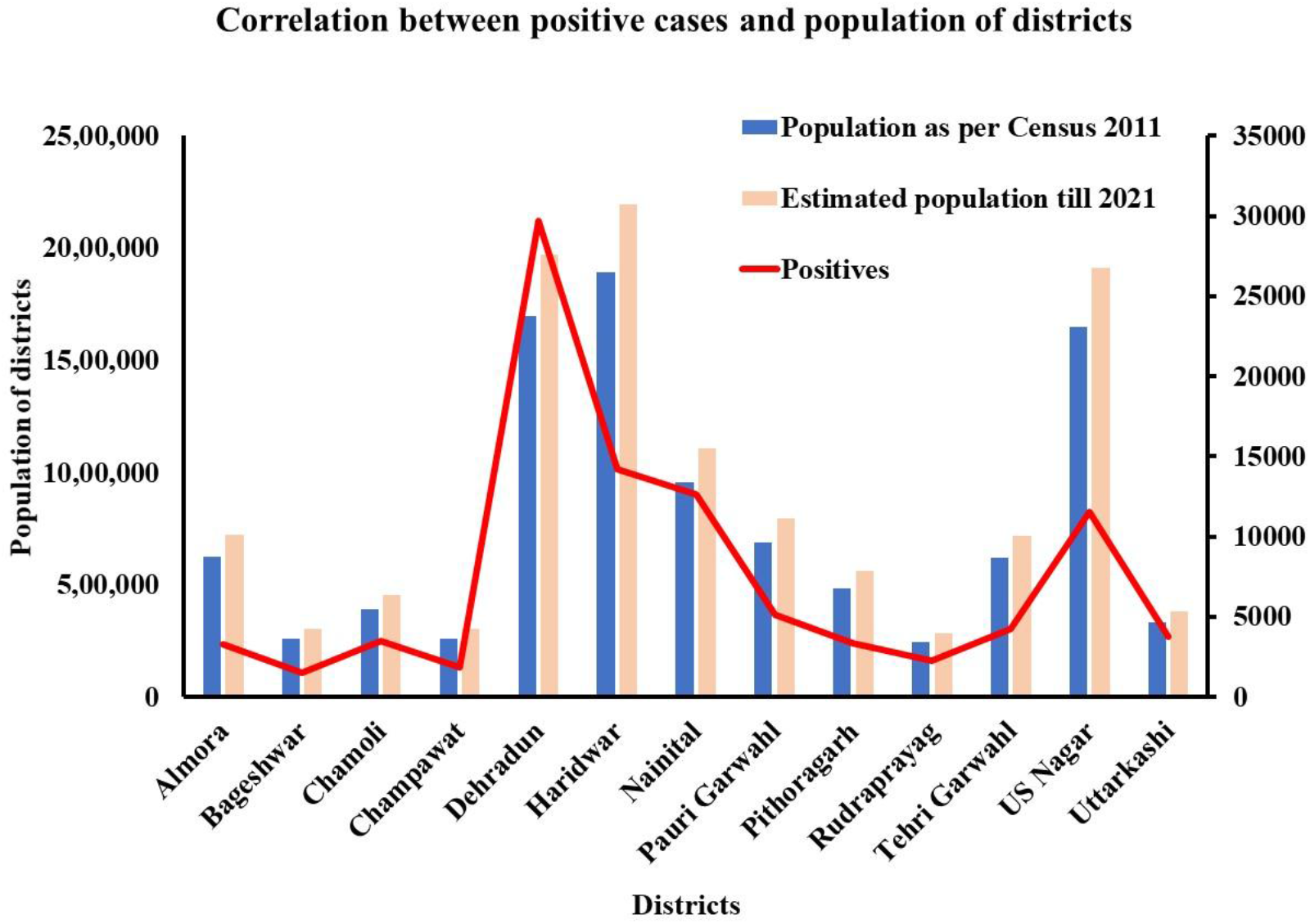
The graph reveals a high correlation between population among 13 districts of Uttarakhand with the outcome of Covid-19 positivity

Further, we attempted to find a correlation between geographical area and prevailing density with the number of Covid-19 positive cases around 13 districts of Uttarakhand. Our analysis (**Figure 7**) revealed a high population density in US Nagar (648), Haridwar (801), Dehradun (550), and Nainital (225) with respect to their respective geographical area in a square kilometer. The high population density with respect to the geographical area is also one of the vital factors for high Covid-19 positive cases in these districts. This has eventually led to the non-implication of social distancing norms which are foremost required in such pandemic situations. The districts like Dehradun, Haridwar, US Nagar, and Nainital were severely affected in contrast to their geographical area, suggesting a high population in these particular districts.

**Figure 7.**
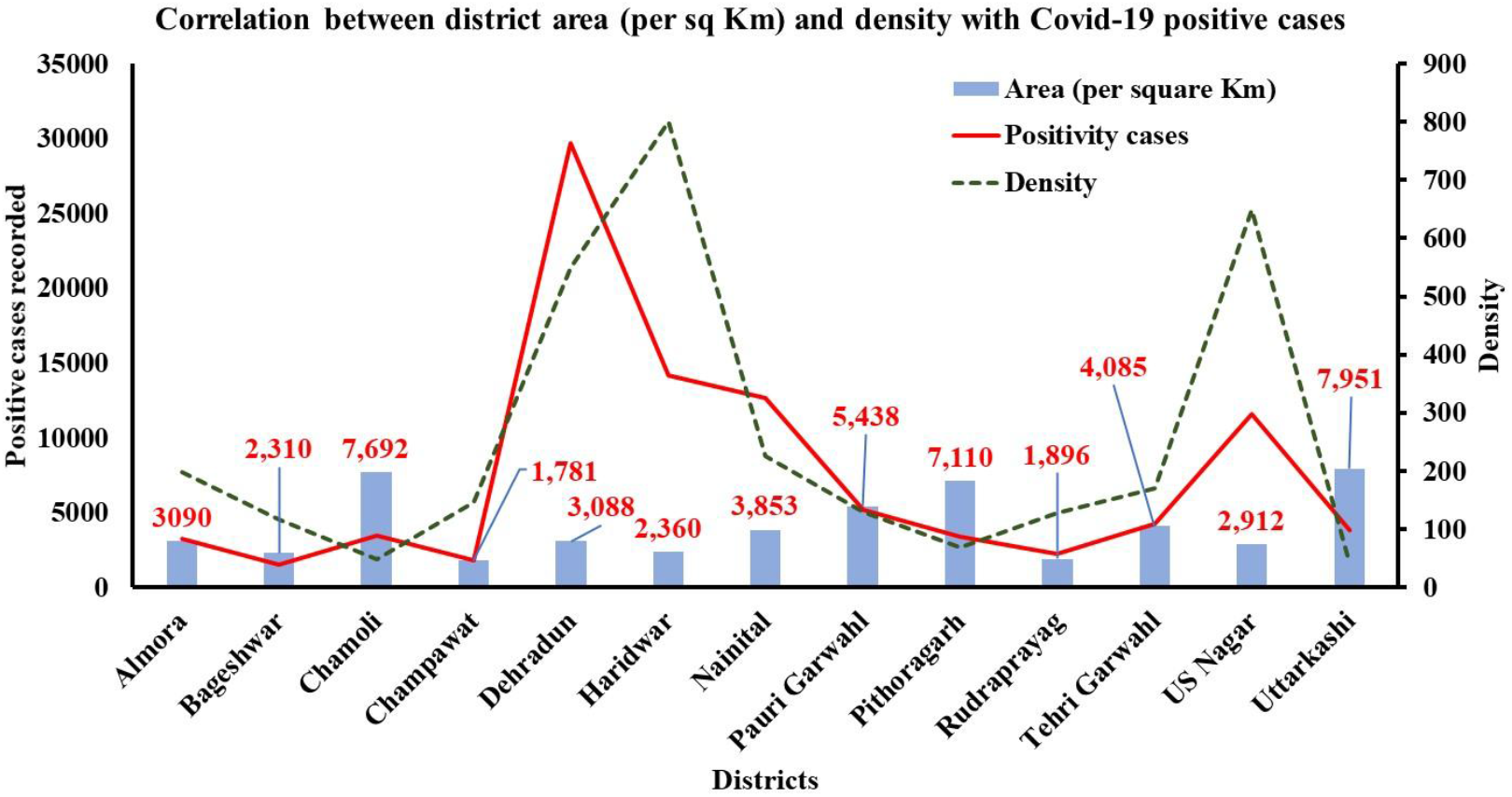
The graph attempts to correlate the total geographical area along with the density of each district of Uttarakhand and its implication of Covid-19 positive cases. The analysis revealed a high positivity rate among districts with high density with a low geographical area

As per Press Information Bureau, latest on February 17, 2021, released a comparative analysis of Covid-19 impact on all States and Union territories of India^8^. As per data, these regions were classified into three groups, *viz*. green zone (1-30000 cases); orange zone (30001-280000 cases); and red zone (above 280000). The Uttarakhand was well placed in the orange zone with 96722 active cases, 94396 recovered cases,and 1678 people deceaseduntil the press release date. An overlook over the data (**Figure 8**) suggested Uttarakhand holds 17^th^ position compared to the severity of Covid-19 among 36 States/Union Territories of India. The worst Covid-19 affected States/Union Territories than Uttarakhand includes (top to bottom), Maharashtra, Tamil Nadu, Karnataka, Delhi, West Bengal, Uttar Pradesh, Andhra Pradesh, Punjab, Gujarat, Kerala, Madhya Pradesh, Chhattisgarh, Haryana, Rajasthan, Jammu and Kashmir, and Odisha. Another similar geographical state, i.e., Himachal Pradesh, was shown to exhibit fewer cases (58142) and deaths (990 cases) in comparison to Uttarakhand during the period of analysis.

**Figure 8.**
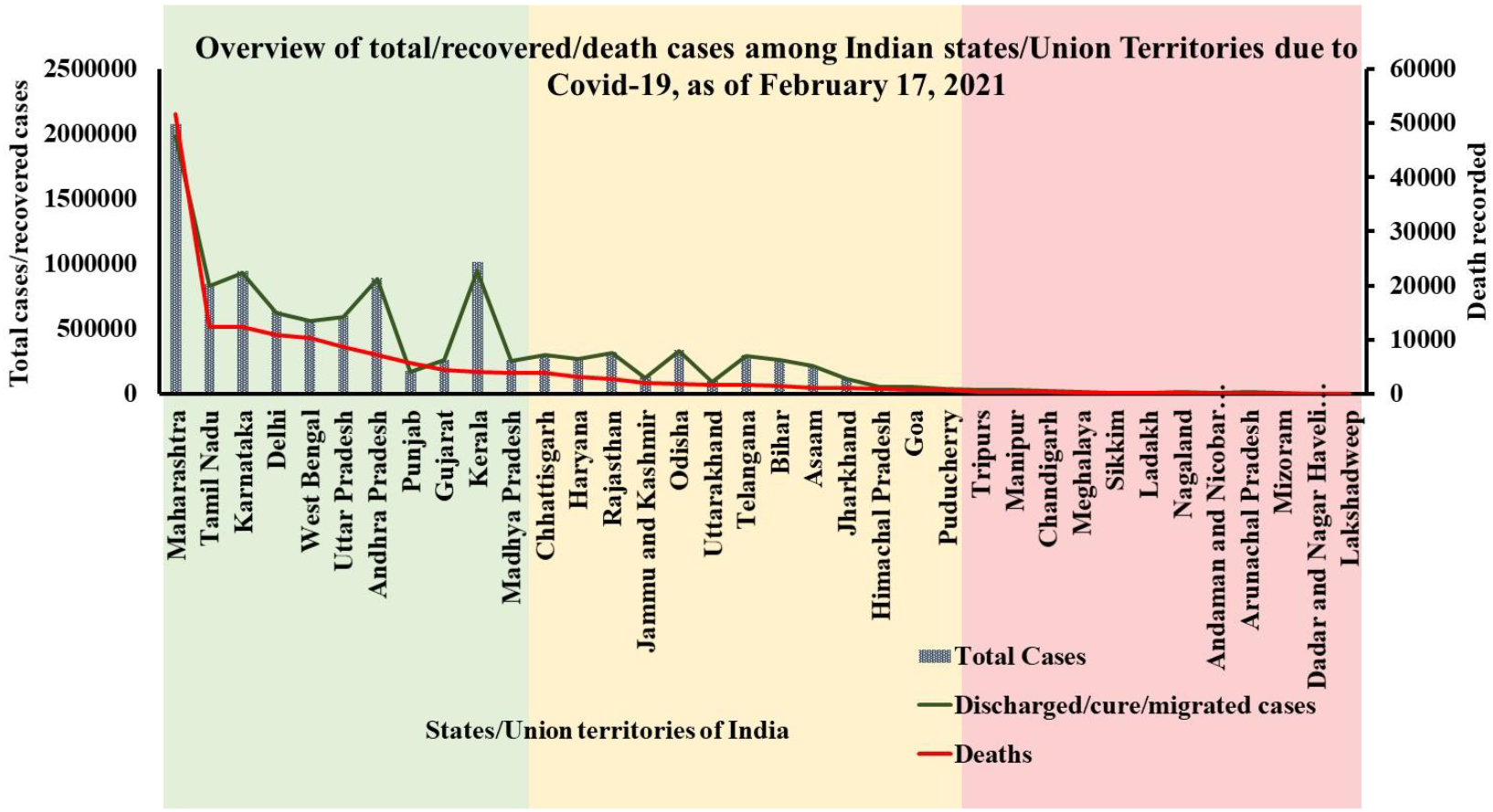
Graphical representation to showcase total/recovered/death cases among India states and Union territories due to Covid-19, as of February 17, 2021

Moreover, we analyzed (**Figure 9**) to decipher positivity, recovery, and death percentages among Indian states/Union Territories due to Covid-19 during the said period of analysis. The analysis revealed that the positivity and recovery percentage exhibit a high correlation among all the states and union territories of India. Maharashtra experienced a high positivity rate of 18.94%. This was followed by Andhra Pradesh (8.12), Tamil Nadu (7.73), Karnataka (8.65%), Delhi (5.82%), and so on. This was further found overlapping with recovery rates, with Maharashtra portraying a recovery rate of 18.62, followed again by Andhra Pradesh (8.28), Tamil Nadu (7.79), Karnataka (8.72%), Delhi (5.87%). Moreover,a high death percentage was also associated with these states, where Maharashtra exhibited a high of 33.10%. This was followed by Tamil Nadu (7.79), Karnataka (7.87%), Delhi (6.99%), West Bengal (6.56%), and so on. Uttarakhand unveiled a positivity percentage of 0.884, recovery percentage of 0.887,and death percentage of 1.076 during the analysis.

**Figure 9.**
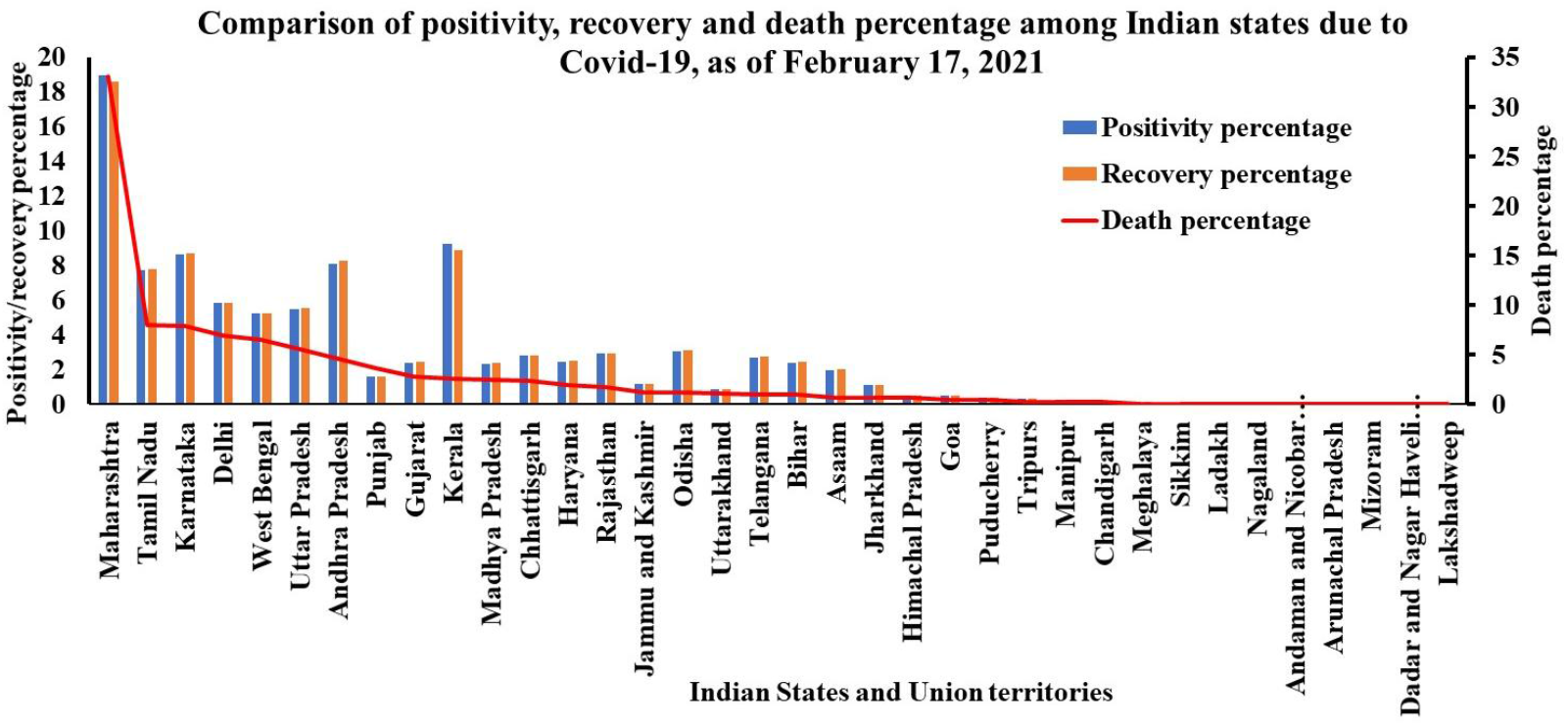
Graph representing positivity, recovery, and death percentages among Indian states/Union Territories due to Covid-19 during the period of analysis

## 4. Future strategies to mitigate such pandemics in future

In the current work, we have analyzedthe impact of Covid-19 in the state of Uttarakhand during its first wave (April 1, 2020, to February 28, 2021, period of analysis).The data generated based on its 13 districts, including Almora, Bageshwar, Chamoli, Champawat, Dehradun, Haridwar, Nainital, Pauri Garhwal, Pithoragarh, Rudraprayag, Tehri Garhwal, U. S. Nagar, and Uttarkashi, revealed a total of 20,70,626 samplesanalyzed among which 97021 were found positive, and 1693 deaths were reported during the period of analysis. Among the districts, Dehradun, Haridwar, Nainital, and US Nagar were found severely impacted by the Covid-19 upsurge in the state and reported peak positive cases during the 21^st^ – 26^th^ week. However, the hilly terrains of Uttarakhand districts, including Chamoli, Pauri Garhwal, and Rudraprayag, reported a high number of positive cases between the 30^th^ and 31^st^ week, which was more in terms of total average throughout the pandemic. In addition, almost every hilly district of Uttarakhand reported an increase in Covid-19 cases during the 34^th^ to 38^th^ week.

Further, the population of a particular district was found to be directly correlated with Covid-19 infective cases. However, this was inversely correlated to the geographical area. Moreover, the major sigh of relief from the current analysis was a 100% correlation between infective cases and recovery cases which have kept mortality under excellent control. We are of the particular opinion that underlying are the plausible reasons.

### 1. Lockdown

Lockdown is one primary parameter strictly imposed during the first wave experienced by the State of Uttarakhand. The strict lockdown around the state ensured a low number of cases and deaths compared to other states. The lockdown also led to the strict following of the health ministry’s guidelines and created a social distance that breaks the infectivity chain^9^. Uttarakhand recorded the highest SARS-CoV-2 growth during the last week of May 2020. We viewed the high doubling rates of infections once the lockdown was slowly uplifted in the state post-May 3, 2020. The current analysis also revealed the importance of lockdown, social distancing, and advisory on Covid-19 impact.

### 2. Seasonal Outbreak

Viral infections follow seasonal outbreaks, for instance, influenza central prominent peak during the winter (January to March) and a minor but significant peak in the post-monsoon season (August to October). Dengue (August-September), and Chickenpox usually after winter season (Jan-April), etc. Last year, the Covid-19 peak was reported on September 19, 2020, in Uttarakhand with 2078 cases in a day, and the second-largest single-day spike in fresh 1946 Covid-19 cases was appeared as on November 19, 2020. At this moment, massive vaccination drive is being carried all over India including Uttarakhand state. Hence, it is also expected that after November 2021, Covid-19 cases may likely decrease. In terms of doubling rate ranges from 4.94-6.70, maximum growth of the virus was obtained in first week of Jun 2020 in Uttarakhand. Here district Bageshwar has experienced the highest doubling rate 6.70 as on 07th Jun, 2020. The year 2021 is also experiencing the same growth pattern and the active cases were reducing day by day after Jun 2021, i.e. 76232 active cases as on 18 May, 43520 active cases as on 26 May, 3471 active cases as on 17 June, 2627 active cases as on 25 June. This evidence from the previous COVID-19 pandemic suggests that exposure to crucial seasonal time may likely increase COVID-19 cases in 2021 as well. Hence, COVID appropriate behavior/activities/initiatives need to strictly follow up till Nov 2021 for effective tackling of Covid-19.

### 3. Rapid urbanization of Uttarakhand

More than half of the Uttarakhand population dwells in villages. The only source of income in the households is agriculture, small handloom industries, or tourism. The educated youth of Uttarakhand do not have enough facilities or job opportunities in his nearby vicinity to meet his and his family’s daily bread^10, 11^. Due to lack of job opportunities and fewer earning sources, there is a high migration rate from villages to cities of Uttarakhand and the different parts of countries for a better livelihood^12^. Uttarakhand’s districts having job opportunities majorly include Dehradun, Haridwar, U. S. Nagar, and Nainital. These were the districts that were worst affected by the Covid-19, and the only plausible reason we explore was high population density inversely impacting the social distances norms during such pandemics aroused by the viral diseases. This is also evident with current analysis that portrays a high positivity rate among districts with high density (US Nagar (648), Haridwar (801), Dehradun (550), and Nainital (225) with respect to their respective geographical area. Secondly, the rise in the cases, particularly in hilly terrains of Uttarakhand districts, is attributed to the movement of natives of Uttarakhand that migrated from their different states where they were initially employed once the restrictions were relaxed in the borders of Uttarakhand state. The local spread has only been confined to the hilly regions^13^. The first wave has seen a high upsurge in unemployment. This will again impact the major big cities of Uttarakhand, equipped with better job opportunities by increasing the population density.

### 4. Healthcare infrastructure and facilities

This hilly, geographically diverse state is considered the most vulnerable in healthcare facilities. The healthcare facilities of hilly districts are very much crippled. The majority of them are unequipped to handle the pandemics such as Covid-19. The lack of health infrastructure of Government aided hospital further adds to the agony^14^. This can be directly visualized because prominent well-to-do’s once infected were airlifted to some well-equipped hospitals of other states^15,16^. This is an exemplary saying that if the Government itself does not have faith in their aided hospitals, what example will they set for their native population. This is a debatable issue, and the Government should look at this unmet fulfilment by investing in improving the infrastructures of all the hospitals in the state. In our personal opinion, state Uttarakhand needs minimum of 2 airlift ambulances, one each for Uttarakhand’s Kumaun and Garhwal covering the entire region.

The one drawback of the Uttarakhand state is that it lacks some major research institutes or scientific centers that indulge in drug discovery or development. The laboratory setups and funding received by Government institutes of higher learning are very meagre to carry out sound translational research, mainly focusing on the bench to bedside discoveries. This will assist in creating awareness and interest in science among youth and create a high intellect workforce having out of box thinking that may lead to some ground-breaking research in the area. Even the state of Uttarakhand, which is rich in terms of Flora, cannot provide a medicinally active and more comprehensive acceptable product to the market so far.

### 5. Social networking and Media

This has done more harm than doing good in current Covid-19 scenarios, particularly in the village areas with limited access to the internet and televisions. The scientific guidance’s are least prioritized over rumors, conspiracy theories, misinformation fuelled by rumors, stigma, and conspiracy theories can have potentially severe inverse implications on public health if prioritized over scientific guidelines^17^. The first wave has shown immense signs of lack of information and awareness about the severity of Covid-19 infections among the state population. The Government here did their best to aware people but overspread rumor was much high compared to the factual news^18^. The high indulgence in self-medication and the outburst of mucormycotic infection recently in the state are key examples. Other examples include the high use of naturally derived nutraceuticals that are further associated with kidney impairment^19,20^.

To address these pertaining issues, immediate action on following suggestions that may curb Covid-19 or even other severe pandemics in the state in the future. We fully understand that lockdown is a haunt for small vendors, Private organization employees, and other daily wagers. This not cripples them economically, mentally, and emotionally but also impacts the development of the country. To plan lockdown more effectively, the **a**. government should focus on the job security of its said class of employees during such pandemics and work on making them financially independent by launching various schemes; **b**. should plan to develop more employment opportunities in hilly terrains and other smaller district, so the youth find competent employment near their dwelling places thus relieving the strain of migration to big cities; **c**. more entrepreneurship modules should be developed with better awareness and start-ups grants to popularise them among youth and prevent their migration.

The health infrastructure is one key area of significant concern, and these should be **a**. equipped with highly sophisticated testing facilities, life-saving devices, quality medicines, among others; **b**. the secondary healthcare workers, including pharmacists, nurses, and other allied persons, should be thoroughly trained and be made prepared for future pandemics. This will immensely reduce the high-end burden on primary health care servants; **c**. high sophisticated laboratories should also be developed in the region to overcome the dependency on other state’s laboratories and timely diagnosis of infection or other diseases; **d**. Adopting telemedicine as aneffective tool to ensure the health facilities in the rural and remote areas of Uttarakhand^21^.

Further, internet connectivity, electricity, and proper roads should be there to connect with the rural areas of Uttarakhand. The people should be provided basic knowledge on ‘digital India’ schemes to make them aware and trained to use the internet in a much better way. The awareness will help curb the myths and rumors that were most prominent during the spread of Covid-19. The better roads will help connect with hospitals easily and in quick time andthus, will be a life-saving effort. Last but not least, the policymakers should work in a much better way to develop some scientific institutes especially concerned with high-end and World-class research in pharmaceutical and biological sciences. The institutes already working in high-thrust areas of health sciences should be funded to carry out the high-end research, which not only in other states but could be recognized internationally. An important parameter of academia-industry collaborations should also be looked upon.

Finally, as we are that Covid-19 is not the last pandemic which the World is witnessing right now. Thus, this is high time to think constructively to mitigate such pandemics in the future with significantly less mortality andsafeguarding economic and socialrequirements. The driving motive behind current work is to correlate the underlying factors and improve preparedness. Moreover, similar studies may assist other states and country(s) re-evaluating the shortcomings of the current pandemic in a much better way and act much better in the future.

## Data Availability

Free for public use

https://health.uk.gov.in

## Acknowledgement

Authors are thankful to Prof. Kamal Ghansala, Chancellor Graphic Era Hill University for providing all necessary facilities. Authors are also thankful to Uttarakhand Government for free access of Covid-19 Uttarakhand data.

## Conflict of interest

The authors declare no conflict of interest.

## Notes

### Competing Interest Statement

The authors have declared no competing interest.

